# Stiffness Matters: Improved Failure Risk Assessment of Ascending Thoracic Aortic Aneurysms

**DOI:** 10.1101/2023.03.09.23287075

**Authors:** Klaas Vander Linden, Emma Vanderveken, Lucas Van Hoof, Lauranne Maes, Heleen Fehervary, Amber Hendrickx, Peter Verbrugghe, Filip Rega, Bart Meuris, Nele Famaey

## Abstract

**Background:** Rupture and dissection are feared complications of ascending thoracic aortic aneurysms caused by mechanical failure of the wall. Currently, the largest aortic diameter is used to predict the risk of wall failure and to determine the need for surgical resection. However, this criterion lacks accuracy, and other prospective parameters are required to predict aneurysm rupture or dissection.

**Methods:** To identify better predictors, we performed a retrospective personalized failure risk analysis, including clinical, geometrical, histological and mechanical data of 33 patients. Uniaxial tensile tests until failure were performed to determine the wall strength. Material parameters were fitted against *ex vivo* planar biaxial data and *in vivo* pressure-diameter relationships at diastole and systole. Using the resulting material properties and *in vivo* data, the maximal *in vivo* stress at systole was calculated, assuming a thin-walled axisymmetric geometry. The retrospective failure risk, defined as the ratio between the maximal stress and maximal strength, was correlated with prospective parameters to find the best failure risk predictor.

**Results:** Our results show that the distensibility coefficient, which reflects aortic compliance and is derived from pressure measurements and multiphasic scans, outperforms other predictors in assessing the risk of aneurysm failure.

**Conclusions:** In a clinical setting, multiphasic CT-scans followed by the calculation of the distensibility coefficient can be of added benefit in patient-specific, clinical decision making. Additionally, the distensibility derived from the aneurysm volume has the best predictive power as it also takes the axial stretch into account.

Clinical trial URL: https://clinicaltrials.gov/ct2/show/NCT03142074?cond=Aneurysm+Ascending+Aorta&cntry=BE&city=Leuven&draw=2&rank=1

**Clinical Perspective:** *What is new?:* The distensibility coefficient, which reflects the capacity of the vessel to expand during pressure changes, outperforms purely geometrical predictors when assessing the rupture or dissection risk of ascending thoracic aortic aneurysms. Not only the change in diameter, but also the change in axial length is important to assess the risk of aneurysm failure. Both dimensions are considered when the change in aneurysm volume is measured.

*What are the clinical implications?:* To quantify the distensibility when determining the need for surgery, the aneurysm should be visualized at two phases, *i*.*e*. at diastole and systole. Therefore, ECG-triggered scans with at least two phases need to be included in clinical practice. Clinicians can add this new metric to their risk stratification process, where it can gradually replace purely geometrical predictors as the database and hence confidence in a cut-off value grows.

## 1. Introduction

An ascending thoracic aortic aneurysm (ATAA) is a chronic degenerative disease characterized by a permanent dilation of the aortic wall, which can culminate in a life-threatening emergency when the wall ruptures or dissects [1, 2]. In theory, prophylactic replacement of the aorta should be performed when the risk of rupture or dissection is greater than the interventional risk [3]. In clinical practice, the rupture or dissection risk is assessed by geometric factors. Currently, surgical repair is recommended when the maximum aortic diameter exceeds the threshold value of 55 mm, or when the growth exceeds 5 mm per year [4]. The mentioned values may vary in case other risk factors are present, such as a bicuspid aortic valve, genetic syndromes (Marfan, Loeys-Dietz and Turner syndrome), hypertension and family history of dissection [5, 6]. Research has shown that the diameter criterion poorly represents the individual risk, resulting in a large number of false positives and false negatives [7, 8].

In search of better predictive tools to assess the failure risk, some have indicated that the diameter should be indexed by patient body surface area (aortic size index, ASI) [9] or height (aortic height index, AHI) [10]. Others have pointed out that the axial length of the ATAA, measured as the distance between the annulus and the brachiocephalic artery, may improve the risk stratification [11].

Essentially, wall failure is a biomechanical phenomenon, since it occurs when the load,*i*.*e*. wall stress or strain, exerted on the aortic wall exceeds its load bearing capacity,*i*.*e*. wall strength or extensibility [12]. Therefore, biomechanical models to predict the failure risk have gained increasing attention. Martin *et al*. compared the aortic wall stiffness, *i*.*e*. with the risk of dissection or rupture [13]. Although a decreased compliance seemed to correlate with an increased risk, one major limitation of this study was that they neglected the *in vivo* wall thickness when calculating the risk, so there was no compelling evidence of the wall stiffness’ risk potential.

Despite these findings, recent publications demonstrate that the ongoing clinical debate is still primarily dominated by the diameter criterion [14, 15, 16]. There is need for clear evidence for more reliable guidelines to identify patients at risk of aneurysm rupture or dissection. To identify reliable & clinically measurable predictors, we performed a retrospective personalized failure risk analysis, including clinical, geometrical, histological and mechanical data from patients undergoing surgery for an ATAA.

This study is structured as follows. Section 2 explains the histological, geometrical and *in vitro* and *in silico* mechanical analysis to estimate the wall strength, stress and failure risk. Section 3 presents the correlation of the failure risk in function of non-invasive predictors. Section 4 discusses the benefit, the consequences, applicability and limitations of the presented results. Finally, Section 5 summarizes the most important conclusions.

## 2. Materials and Methods

The study was approved by the Ethics Committee Research UZ/KU Leuven (NCT03142074) and patient informed consent was obtained. Patients with genetic syndromes such as Marfan, Loeys-Dietz and Turner, were excluded from the study. The study included 33 patients who underwent surgery at the University Hospitals Leuven. Prior to surgery, patients underwent a ECG-gated contrast-enhanced CT scan and the diastolic (*p*_*dias*_) and systolic (*p*_*sys*_) pressures were measured non-invasively. All patients were operated under cardiopulmonary bypass with arrested heart via sternotomy. The ascending aorta was excised and sent for further analysis.

### 2.1. Histological analysis

Samples for histological analysis were fixed in paraformaldehyde (6%), dehydrated (Medite TES 99) and embedded in paraffin. 5-μm-thick serial cross-sections were created (Microm HM360) and stained with Hematoxylin and Eosin and Elastica van Gieson stains using standard laboratory protocols. All specimens were microscopically examined (Philips Ultra Fast Scanner). The thickness of the entire wall and sublayers, *i.e*. intima (I), media (M) and adventitia (A), were calculated as the average at ten different locations of the slice with ImageJ 1.52a (National Institutes of Health, US). The fractions of the wall constituents, *i.e*. elastin, collagen, smooth muscle cells, were measured using an in-house developed image processing software written in Matlab R2019a (Mathworks Inc., US). Presence of intimal hyperplasia and medial degeneration (moderate or severe) was qualitatively assessed according to guidelines presented in [17]. All measurements and assessments were performed by two independent observers.

### 2.2 Geometrical analysis^l^

Preoperative ECG-gated CT scans, each consisting of approximately 600-800 transversal slices with an in-plane resolution of 0.34 mm and a slice thickness of 0.6 mm, were segmented and reconstructed into a 3D model at end-diastole (*k* = *dias*, 0% RR-interval) and mid-systole (*k* = *sys*, 33% RR-interval) phase using Mimics Innovation Suite 23 (Materialise, Belgium) [18]. At both phases *k*, the luminal volume of the aorta was segmented, see Figure 1. The maximal diameter *D*_*k*_ was defined by the best-fit circle along the center line (CL) at the widest point of the aneurysm. The length of the aneurysm *L*_*k*_ was defined as the distance along the CL between the sinotubular junction and the first bifurcation, *i.e*. the brachiocephalic artery. The volume of the ATAA *V*_*k*_ was defined as the volume of the aorta measured along the length *L*_*k*_. Moreover, *AHI* was calculated as 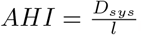 with *l* the height of the patient [10] and the *ASI* was calculated as 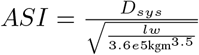 with *w* the weight of the patient [19].

**Figure 1:**
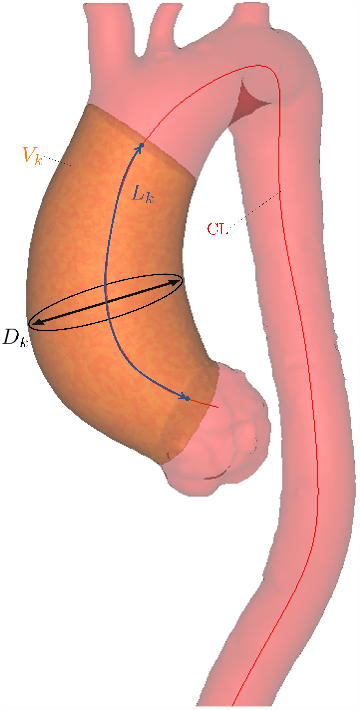
Example of a segmented aorta (pink) with the ATAA (orange) and CL (red). The aneurysm’s volume (*V*_*k*_), maximal diameter (*D*_*k*_) and length (*L*_*k*_) were measured based on the segmentation at phase *k* = *dias, sys*.

### 2.3. Mechanical analysis

#### 2.3.1. In vivo mechanical characterization

The *in vivo* distensibility coefficient (DC) defines the vessel compliance and reflects the capacity of the vessel to expand during pressure changes. The DC based on the cross-sectional area is defined as [20, 21]:

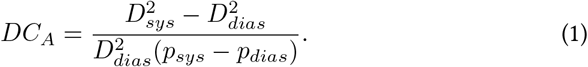

Equation 1 ignores the geometrical changes in axial direction of the ATAA between diastole and systole. Therefore, we define an alternative measure for the DC, based on the volume:

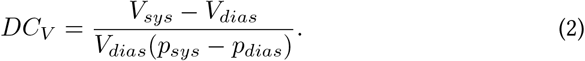

#### 2.3.2. In vitro mechanical characterization

After aortic replacement, the excised tissue intended for mechanical testing was preserved in phosphate-buffered saline solution at -80°C. Experiments were performed at FIBEr, KU Leuven Core Facility for Biomechanical Experimentation. After thawing overnight at 4°C, the tissue was cut open axially and the samples, hourglass-shaped for uniaxial or square-shaped for biaxial tensile testing, were excised according to the tissue’s circumferential (*θ*) and axial (*z*) direction of the aorta, see Figure 2. In some cases, insufficient tissue was available for testing all the indicated samples. Sample thickness was measured with a VHX-6000 Digital Microscope (Keyence Corporation, Japan). All tensile tests were performed using a Messphysik testing device (ZwickRoell, Germany). Images were captured at 20 Hz with Vic-Snap (Correlated Solutions, US, with isi-sys, Germany, as system integrator) using a Manta G-917B camera with a Sony ICX814 monochrome charged-coupled device, mounted perpendicularly to the sample’s surface. The samples were submerged in saline solution at 37°C during mechanical testing. For a detailed description of the test protocols, the reader is referred to Appendix A.

**Figure 2:**
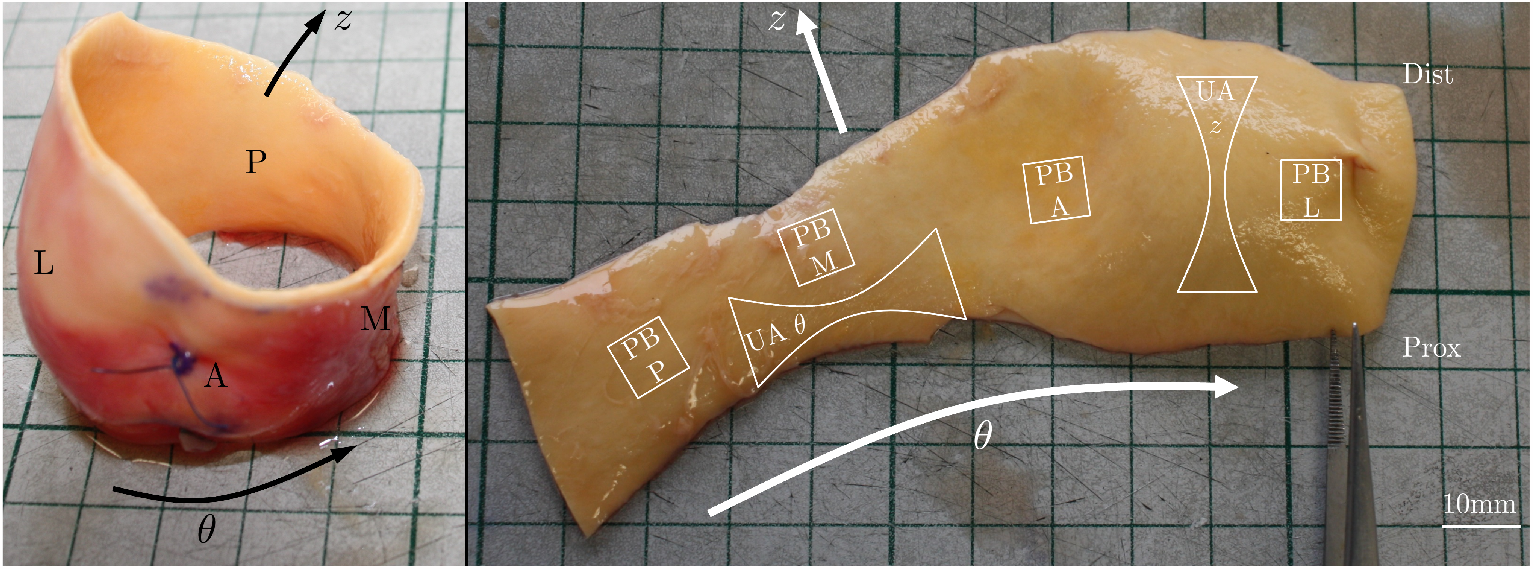
Excised, load-free (left) and opened, stress-free (right) ATAA. Hourglass and square-shaped samples for uniaxial (UA) and planar biaxial (PB) tensile testing are indicated, respectively, as well as the proximal (prox) and distal (dist) side, and the axial (*z*) and circumferential (*θ*) direction. PB samples were extracted from the four quadrants (A: anterior; M: medial or inner curvature; P: posterior; L: lateral or outer curvature).

To estimate the wall strength, hourglass samples were mounted on a uniaxial (UA) test set-up using clamps and were loaded until rupture, see Figure 3. The strength of the tissue 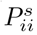 in the direction *i* = *θ, z* is expressed in terms of the first Piola-Kirchhof stress and is calculated as:

**Figure 3:**
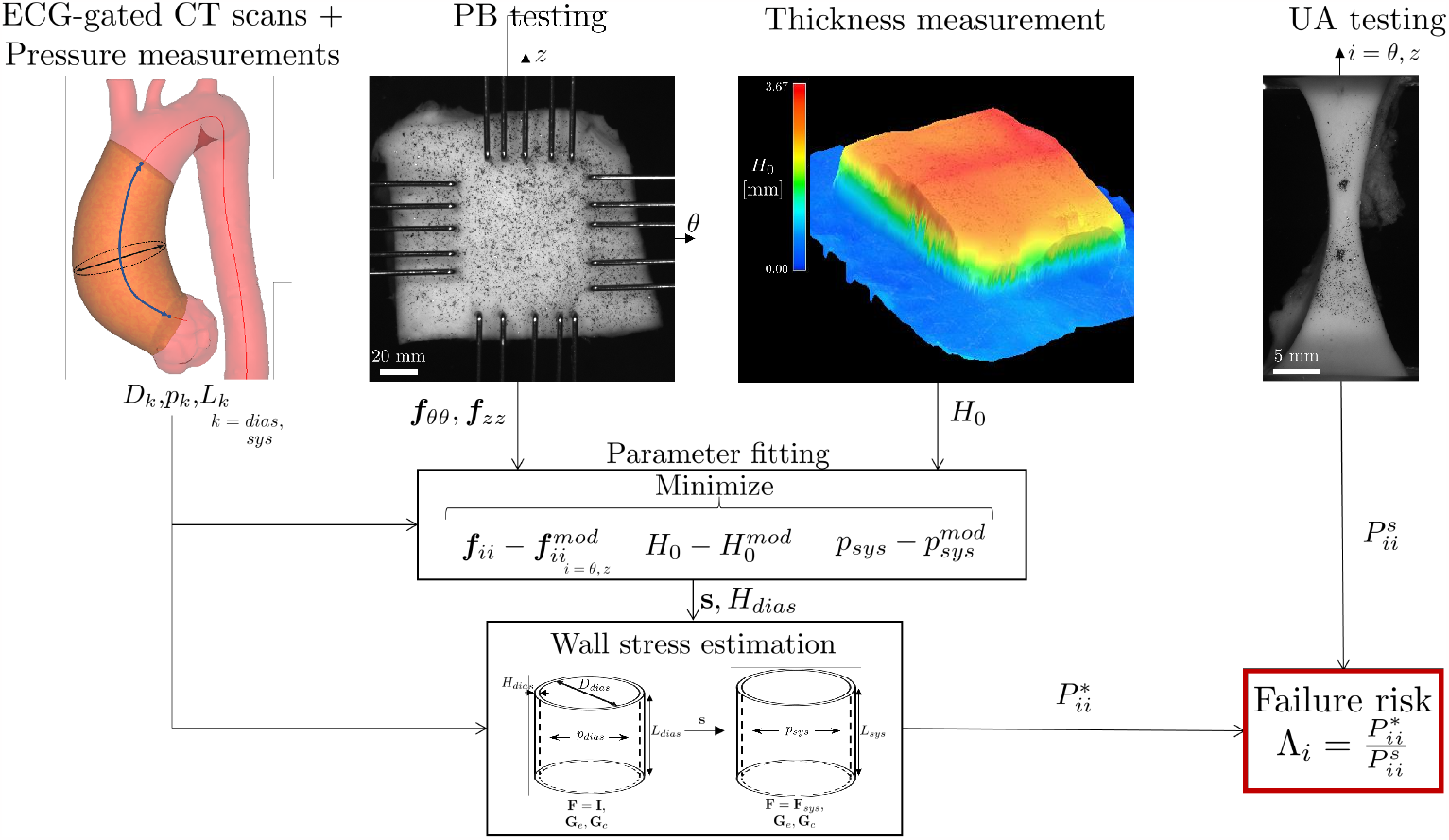
Overview of the retrospective personalized failure risk assessment.

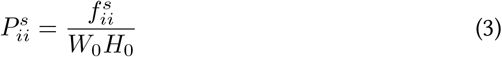

with the normal 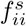 force measured at yield, defined according to [22], in direction *i*, and *W*_0_ and *H*_0_ the width and thickness measured at the gauge section in the stress-free state, respectively.

To characterize the mechanical behavior, square samples were mounted on a planar biaxial (PB) testing set-up using rakes, see Figure 3. The samples were cyclically loaded along their circumferential and axial axes. Experimental forces in the circumferential (*f*_*θθ*_) and longitudinal direction (*f*_*zz*_) were measured. The corresponding deformation at the center of the sample was measured with digital image correlation (DIC).

### 2.4. Determining failure risk

Calculation of the intraluminal maximal wall stress requires information on the material behavior and *in vivo* wall thickness of the tissue. The material parameters **s** and wall thickness at diastole *H*_*dias*_ were estimated using a material fitting approach explained in [23, 24], by minimizing the difference between (1) the experimental (*f*_*ii*_) and model 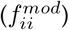 reaction forces in the direction *i* = *θ, z* of the planar biaxial test, (2) the measured (*H*_0_) and model 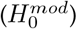 *ex vivo* thickness and (3) the measured (*p*_*sys*_) and model 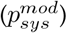 systolic blood pressure, see Figure 3. Knowing the material parameters and *in vivo* thickness, the maximal first Piola-Kirchhoff wall stress at systole 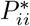 was calculated while approximating the aneurysm as a thin-walled cylinder. More specifically, the aneurysm at diastole was further pressurized and axially stretched using *p*_*sys*_ and 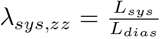, respectively. For a detailed description of parameter fitting approach and the maximal *in vivo* stress calculation, the reader is referred to Appendix B.

The failure risk *Λ*_*i*_ in the direction *i* is calculated as the ratio between the maximal estimated *in vivo* wall stress and the wall strength: 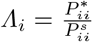. A schematic represen-tation of the failure risk estimation is shown in Figure 3. A low value of *Λ*_*i*_ means that the aneurysm is unlikely to rupture or dissect in the direction *i*, and *vice versa*.

### 2.5. Statistical analysis

Statistical analysis was performed using RStudio. Comparisons were determined using the Mann-Whitney U test and the Fisher’s exact test for continuous and categorical variables, respectively. Correlations were determined using the Spearman rank non-parameteric test. A p-value lower than 0.05 was considered statistically significant, a p-value lower than 0.01 was considered highly statistically significant. Simple logistic regression was performed to establish a relationship between *Λ*_*i*_ and possible predictors.

## 3. Results

### 3.1. Subject characteristics

The subject characteristics are summarized in Table 1. The data set was divided into two different groups depending on their volume-based distensibility coefficient *DC*_*V*_ in relation to 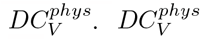 has been reported to be 0.25*e*-2mmHg^-1^[20]. If 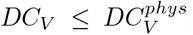, the aneurysm was classified as ‘stiff’, if 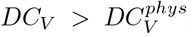, the aneurysm was classified as ‘compliant’. Figure 4 shows the total systolic volume, pressure and volume-based distensibility in function of the patient’s age.

**Table 1:**
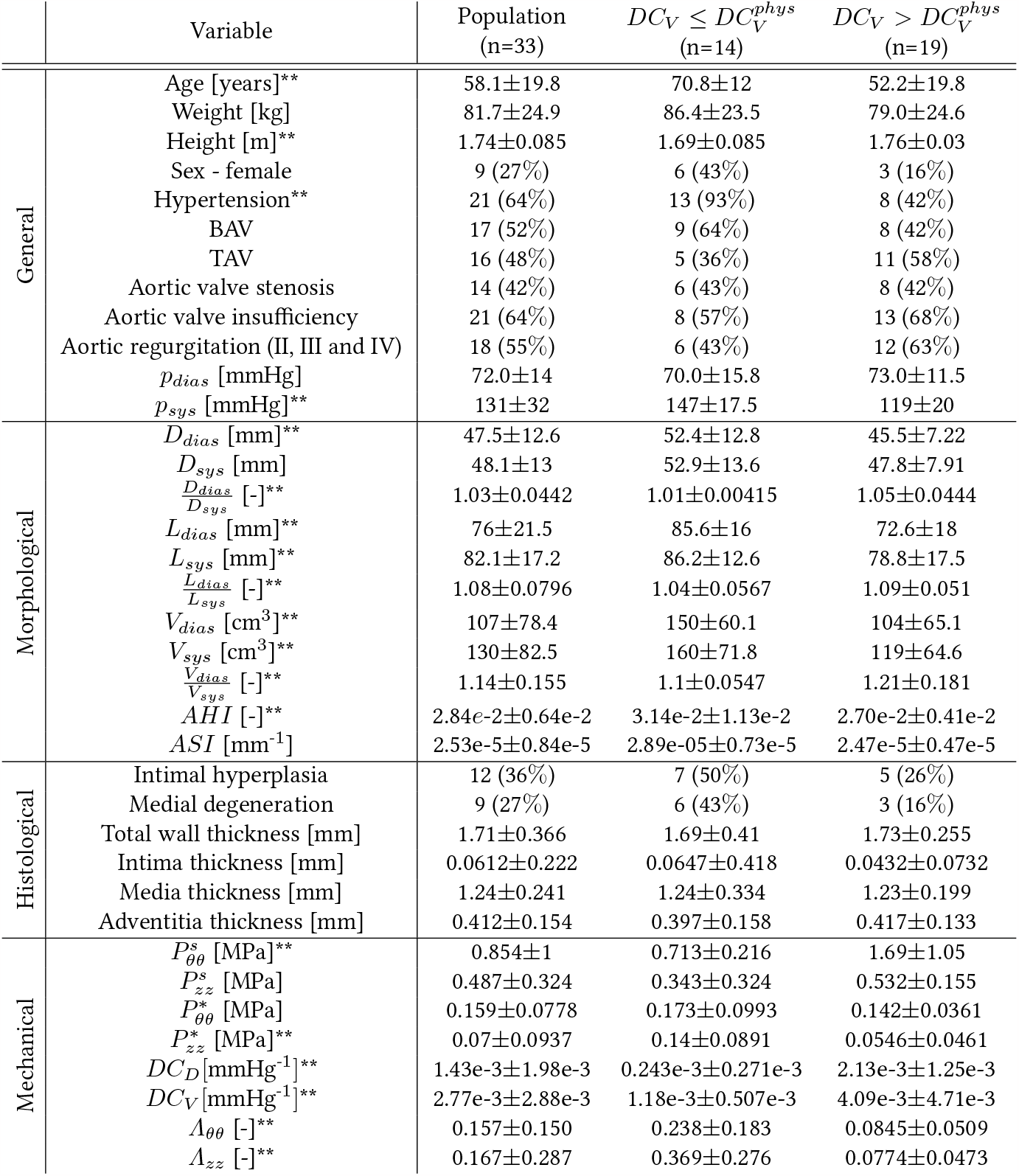
Baseline characteristics. Results are shown as median ± interquartile range or as number of patients (percentage). *Ddias* and *Dsys* refer to the maximal aortic diameter measured at diastole and systole, respectively. Aortic regurgitation was classified according to the guidelines presented in [41]. A p-value < 0.05 is indicated with *. A p-value < 0.01 is indicated with **.

**Figure 4:**
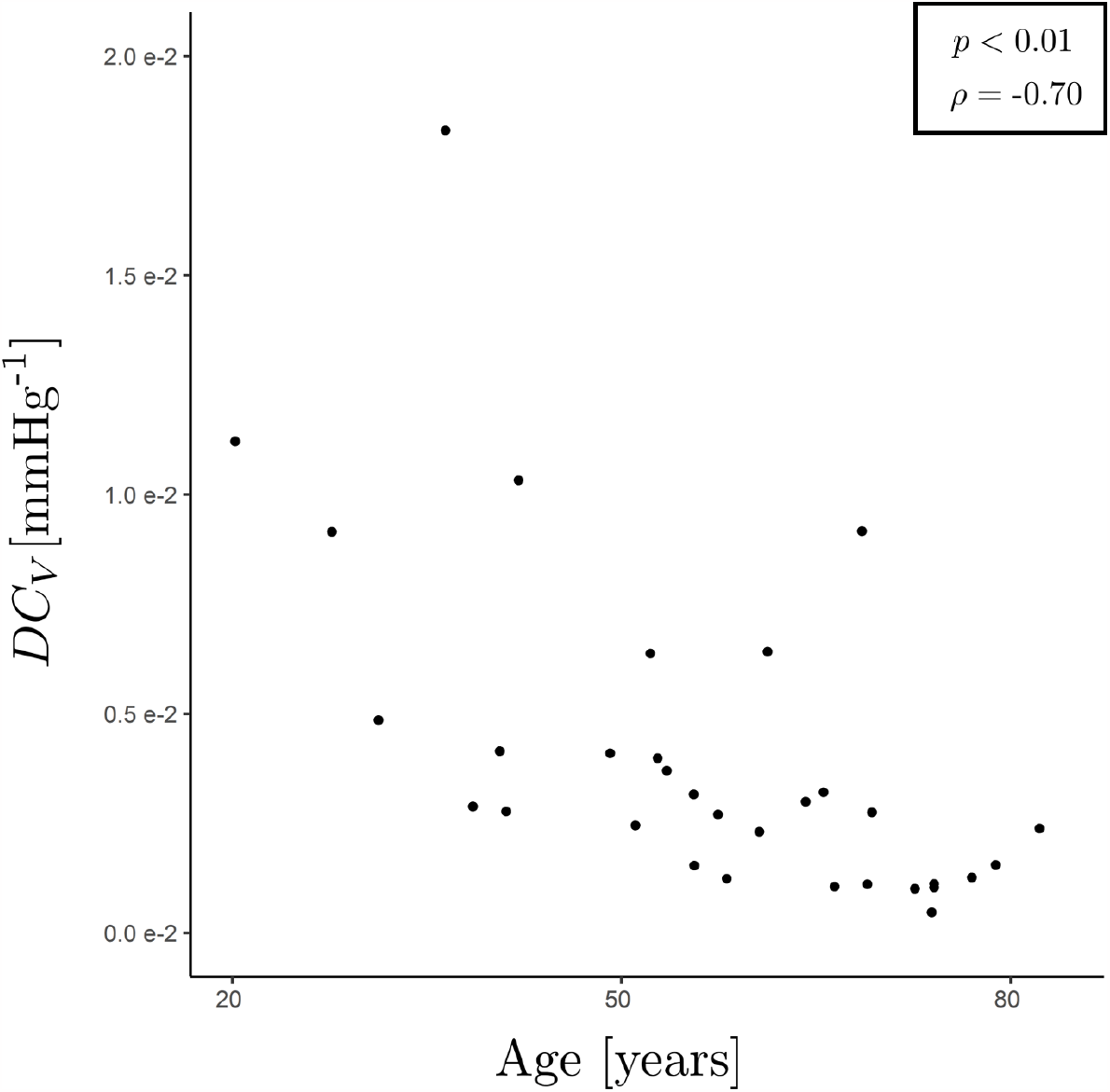
Volume-based distensibility (*DC*_*V*_) in function of age.

The fitted material parameters **s** and diastolic wall thickness *H*_*dias*_ of each patient, together with the fitting measures are shown in Appendix C, Table 4.

### 3.2. Failure risk assessment

Table 2 presents the Spearman *ρ* correlation coefficients with corresponding p-values between the different clinically accessible characteristics and the failure risk *Λ*_*i*_ (*i* = *θ, z*). Figure 5 shows the failure risk *Λ*_*i*_ (*i* = *θ, z*) as function of *D*_*sys*_, *AHI, DC*_*A*_ and *DC*_*V*_. Spearman *ρ* correlation coefficient with corresponding p-value and Nagelkerke pseudo *R*^2^ value of the logistic regression are also shown on the figure. All but one aneurysm was predicted not to rupture or dissect, *i.e. Λ*_*ii*_ *<* 1, *i* = *θ, z*. For the one aneurysm for which *Λ*_*θθ*_ = 1.43 and *Λ*_*zz*_ = 1.39, the failure risk was retrospectively set to 1, since no ATAA had ruptured or dissected prior to surgery. Additionally, a significant increase of *Λ*_*θθ*_ was observed for hypertensive patients. No significant difference could be observed for the failure risk between TAV and BAV patients.

**Table 2:**
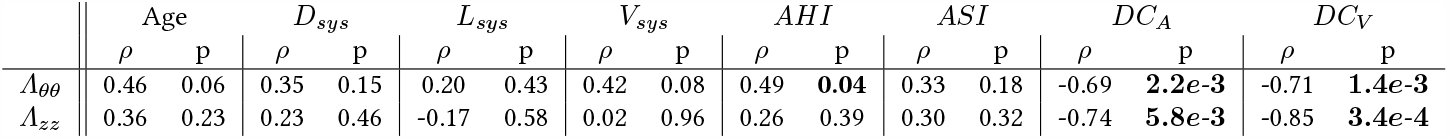
Spearman coefficients with corresponding p-values of the correlation between failure risk and clinically accessible characteristics. Significant p-values are shown in bold.

**Figure 5:**
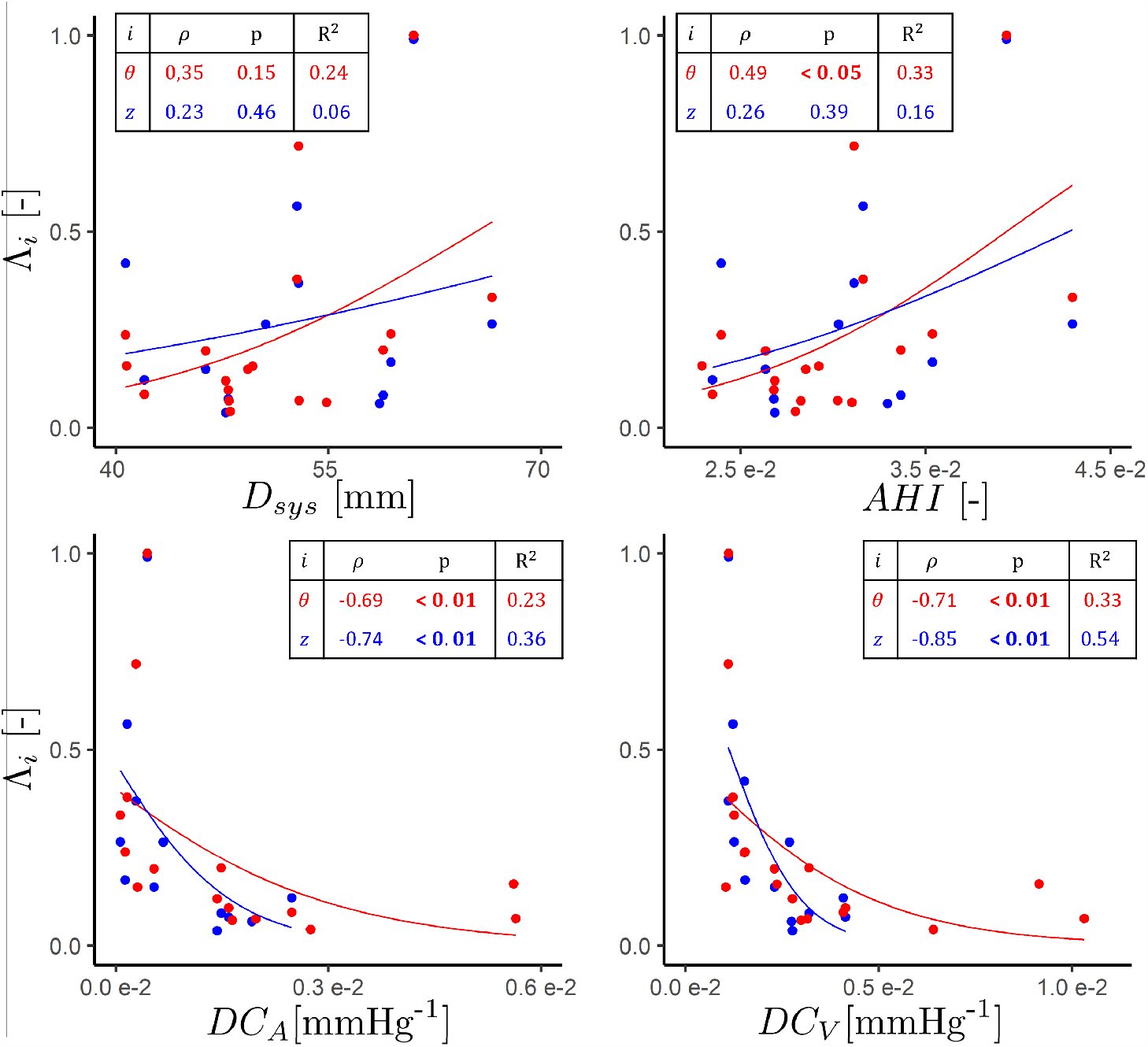
Failure risk in the circumferential (red) and axial direction (blue) with respect to *D*^*sys*^ (top left), *AHI* (top right), *DC*_*A*_ (bottom left) and *DC*_*V*_ (bottom right). For each of the predictors in both directions, the Spearman correlation coefficient with corresponding p-value is given, together with the Nagelkerke pseudo *R*^2^ value of the logistic regression.

## 4. Discussion

We performed a retrospective personalized failure risk analysis of ascending thoracic aortic aneurysms, including clinical, geometrical, histological and mechanical data. Uniaxial tensile tests were performed to determine the wall strength and material parameters were fitted against *in vitro* planar biaxial data and *in vivo* pressure-diameter relationships at diastole and systole. Using the resulting material properties and *in vivo* data, the maximal *in vivo* stress at systole was calculated, assuming a thin-walled axisymmetric geometry. A personalized failure risk was defined as the ratio between the maximal wall stress to the wall strength and compared with clinically accessible predictors in order to predict aneurysm failure. In addition, age-related changes to the aneurysm behavior and wall morphology were investigated.

### 4.1. Aneurysm failure risk assessment

The presented results in Figure 5 and Table 2 confirm once more the inadequacy of the diameter when assessing the failure risk of ATAA [7, 9]. Likewise, alternative geometrical features such as the aneurysm length or volume do not correlate with the failure risk, although they are recommended by others [11]. While the diameter indexed against the patient height (AHI) slightly improves the risk assessment in the circumferential direction, the AHI and ASI (indexing the diameter against the patient body surface area) do not result in statistically significant correlations, contrary to what has been suggested in literature [9, 10, 25]. Furthermore, hypertension, which has been identified as a risk factor in multiple cardiovascular diseases, also led to a higher failure risk in the circumferential direction, [13, 20, 26, 27, 28].

As aneurysm rupture or dissection is primarily a mechanical event, mechanical predictors have the possibility to improve the risk stratification. The distensibility coefficient expresses the ability of the artery to dilate in response to an increase in pressure [20, 21]. The area-based *DC* expresses the compliance in terms of the diameter and relates it to the corresponding pressure increase. The failure risk in both circumferential and axial direction strongly correlates with *DC*_*A*_. The lower its compliance, the stiffer the material becomes and the higher the failure risk. This is in accordance with Martin *et al*. who defined a similar empirical mechanical coefficient, *i.e*. the (diameter-based) pressure-strain modulus to quantify the stiffness of the aorta and found that this predictor has a strong relation with the failure risk [13]. Considering the elevated circumferential stiffness of aneurysm and the axial displacement of the aortic root, the diameter may lower between diastole and systole, which explains why several patients have a negative *DC*_*A*_, see Figure 5. As pointed out by others, the axial dimension is an often overlooked parameter [11]. It is not possible to relate the axial stretch to any kind of measurable axial load. Therefore, the volume-based distensibility *DC*_*V*_ was defined and relates the pressure increase, the only measurable information on the load exerted on the tissue, to the total volume change of the aneurysm. As can be seen on Figure 5 and Table 2, *DC*_*V*_ correlates even better with the failure risk in both directions as compared with *DC*_*A*_. Based on their *DC*_*V*_, patients were divided into two groups, see Table 1. A low *DC*_*V*_, *i.e*. high arterial stiffness, is found in patients with hypertension and with large aneurysms. A low *DC*_*V*_ is also associated with weaker tissue in the circumferential direction and higher stress in the axial direction.

### 4.2. Implications for surgical guidelines

A direct implication of the presented results is the recommendation to visualize the aorta at least at diastole and systole. Only by correlating the deformation between these two phases with the measured luminal pressures can the aortic compliance be assessed *in vivo*. Therefore, we recommend incorporating multiphasic scans into clinical practice when diagnosing aneurysm patients. In addition, clinical practice should not only focus on the circumferential changes of the aneurysm, *i.e*. related to the diameter, but also on axial and volumetric changes. Moreover, the distensibility needs to be measured consistently and accurately. This requires the use of dedicated software to segment the aorta at the time points of interest. The pressures (typically at diastole and systole) must correspond to the appropriate phases at which the aneurysm is segmented and consistently measured [18, 29].

According to Figure 4, the distensibility decreases with age which is in accordance with [20]. Therefore, age seems to have an indirect effect on the failure risk due to the corresponding arterial remodeling and stiffening. Depending on their distensibility, older patients may be at higher risk for rupture or dissection and be more eligible for aortic repair. In this regard, it is important to note that Zierer *et al*. found that elective repair in patients of advanced age, *i.e*. older than 70 years, does not impair functional recovery [30].

### 4.3. Limitations and future work

The *R*^2^ values describing the simple logistic regression between the failure risk and the distensibility coefficients were low to moderate. Additionally, multivariate logistic regression analysis found no significant combinations of predictors that help to predict the failure risk. Therefore, it is not yet possible to determine with sufficient confidence the critical range of predictor values that distinguish stable aneurysms from aneurysms that are likely to rupture or dissect. To do so, more patients are required in the clinical study. The limited data set may also explain why no significant correlations were found between the distensibility coefficient and other cardiovascular characteristics such aortic valve stenosis, insufficiency and regurgitation, see Table 1.

The next crucial step is to build a probabilistic rather than a deterministic framework to assess the failure risk [31, 32]. This includes but is not limited to the usage of stochastic material parameters [33], the uncertainty related to the *in vivo* thickness and wall strength [34, 35] and temporal variations of the blood pressure [27]. Including multiple sources of uncertainty will provide more insight into the relative importance of the input parameters and their influence on the predictors and on the final (probabilistic) failure risk.

We chose for uniaxial tensile testing to estimate the strength of the tissue. Whereas other measurement techniques exist [12], uniaxial tensile testing is a widely accepted method to estimate the strength of the tissue [36]. The benefit of hourglass-shaped samples is that tear will more likely occur at the center of the sample instead of the regions near the clamps due to stress concentrations caused by the test design. The downside of this shape is that deformation at the gauge region is highly inhomogeneous. As a consequence, we were only able to express the failure risk in terms of the (nominal) stress, but not in terms of the deformation [37].

Although we measured the *ex vivo* thickness and mechanical behavior of different tissue sample along the aneurysm circumference, we fitted a single material model to the experimental results. Moreover, we could excise only up to two samples for uniaxial testing (one for each principal direction), and thus did not explicitly account for tissue heterogeneity [38]. We modeled the aneurysms as an axisymmetric homogeneous thin-walled cylinder. The influence of other irregular geometric features such as arterial tortuosity [39] and spatial-dependent material characteristics can be modeled using finite element modeling, but did not fall within the scope of the presented study [40].

## 5. Conclusion

By conducting a retrospective personalized failure risk assessment of ATAA patients, we were able to identify and compare clinically accessible aneurysm failure risk predictors. Mechanical predictors, *i.e*. the distensibility coefficients, outperform predictors based on geometrical features alone in predicting wall failure. The volume-based DC has the best predictive power as it also takes the axial stretch into account. This study shows clear evidence to include multiphasic scans in clinical practice when assessing the risk of aneurysm failure. Clinicians can add this new metric to their risk stratification process, where it can gradually replace purely geometrical predictors as the database and hence confidence in a cut-off value grows.

## Data Availability

The dataset presented in this study is publicly available in KU Leuven RDR.

## Acknowledgements

The authors would like to thank Liselotte Peeters for her help in performing and processing the mechanical experiments, Kimberly Crevits for her help in performing the mechanical experiments and Ruben Snoeck for his help in performing the histological analysis and the segmentation of the CT scans.

## Sources of funding

The authors would like to thank Research Foundation – Flanders (FWO) (Grants no: SB1SA9119N, SB1S56317) and KU Leuven (C2-ADAPT) for providing financial support to this project.

## Disclosures

All authors declare that they have no conflicts of interest.

## Supplemental material

Appendix A: Experimental protocols of the uniaxial and planar biaxial tensile test

### Uniaxial tensile test

Hourglass samples were mounted on a uniaxial test set-up using clamps. The clamp-to-clamp separation at the start of the uniaxial test was 30mm. The protocol consisted of multiple displacement-controlled loading steps: 5%, 10%, 15%, 20%, 25%, 50% nominal strain and until rupture, applied at a linear rate of 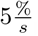. Every loading step consisted of five loading cycles to precondition the tissue. A preload of 0.05 N was applied to flatten the sample. Two markers were applied at the gauge region to track the deformation.

### Planar biaxial tensile test

Square samples were mounted on a planar biaxial (PB) testing set-up using rakes. The rakes consisted of four sets of five parallel needles, each rake spaced 1 mm apart. The sample’s circumferential and axial axes were aligned with the horizontal and vertical testing axes, respectively. The rake-to-rake separation at the start of the test was 7 mm. A displacement-controlled loading protocol was used consisting of six loading steps: 5%, 10%, 15%, 20%, 25% and 35% nominal strain, applied at a linear rate of 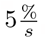. Each loading step was repeated three times, while changing the relative applied loading between the circumferential and axial direction: 1:1; 2:1 and 1:2. Each loading step consisted of four preconditioning cycles and one loading cycle used for further analysis. Before every loading cycle, a preload of 0.05N was applied in both directions to avoid buckling. Using graphite powder, a speckle pattern was applied to the biaxial samples to track the deformation using digital image correlation (DIC). Incremental DIC was performed using VIC-2D (Correlated Solutions, US) with a subset of 97 pixels and a step size of 7 pixels. The deformation gradient throughout the experiment was calculated based on the DIC results of the central 25% of the area enclosed by the rakes, assuming homogeneous deformation [42]. Experimental forces in the circumferential (*f*_*θθ*_) and longitudinal direction (*f*_*zz*_) were captured at 20 Hz and calculated as the average force measured by the horizontally and vertically opposed actuators, respectively. For every ratio, the fifth loading cycle of the last loading step was processed.

Appendix

B: Parameter optimization

### Material description

The Cauchy model stress *σ*^*mod*^ stress is calculated as follows:

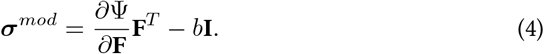

In equation 4, Ψ represents the strain energy of the described material, **F** the total deformation with respect to a reference state, **I** the identity matrix and *b* a Lagrange multiplier. The stresses (both *in vivo* and *in vitro*) are calculated using the plane stress assumption, *i.e*. the stress in the radial direction *r* equals zero: 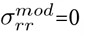, which deter-mines the value of *b*.

The Gasser-Ogden-Holzapfel [43] constitutive material model in the context of the constrained mixture theory is used to calculate the strain energy, based on [23]. The aneurysmal tissue is modeled as a mixture existing of two constituents: the non-collageneous isotropic matrix (*e*) and collagen fibers (*c*). The strain energy function is described as:

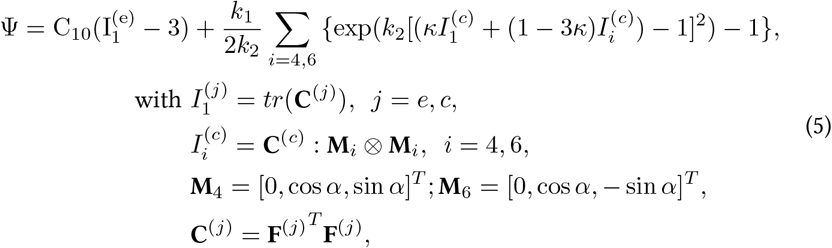

and with **F**^(*j*)^ = **FG**^(*j*)^ representing the elastic deformation experienced by constituent *j* = *e, c*, which consist of the total deformation of the mixture w.r.t. the diastolic reference configuration (**F**) and the constituent specific deposition stretch tensor (**G**^(*j*)^). Figure 6 shows an overview of the different loading states. The parameter set **s** contains the five material properties [*C*_10_, *k*_1_, *k*_2_, *κ, α*], representing the matrix’ stiffness, fiber stiffness, fiber stiffening, fiber dispersion and fiber angle w.r.t. circumferential direction, respectively. The deposition stretch of collagen **G**^(*c*)^ is assumed to be known [44]:

**Figure 6:**
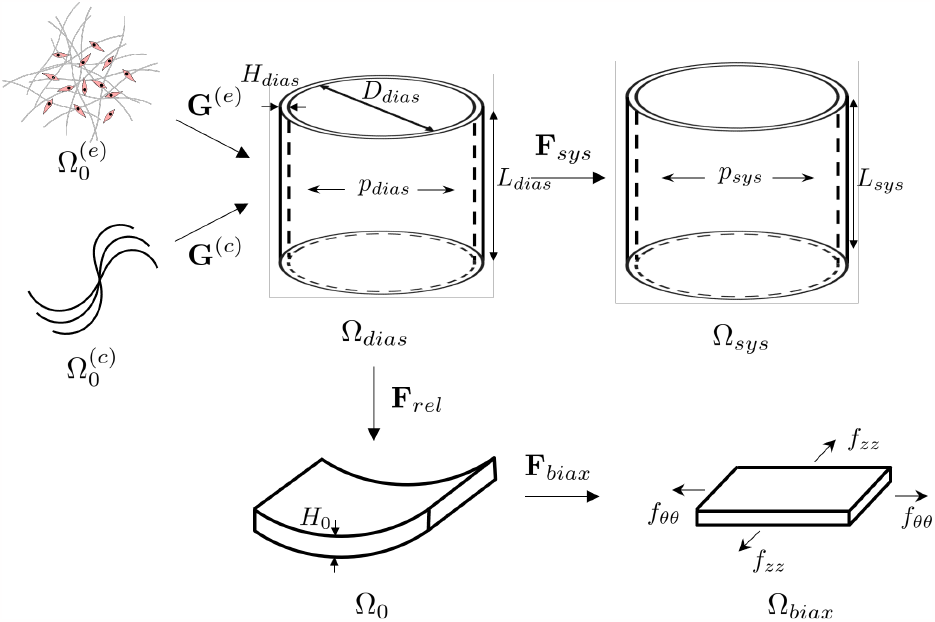
Different loading configurations *Ω* considered during the parameter fitting approach. *Ω*_*dias*_ and *Ω*^*sys*^ represent the *in vivo* diastolic (reference) and systolic configuration, respectively. 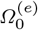 and 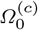 represent the individual stress free state of the non-collageneous isotropic matrix and collagen, respectively. *Ω*_*rel*_ and *Ω*_*biax*_ represents the *ex vivo* zero-stress and biaxially loaded configuration, respectively. Figure partially adapted from [46]

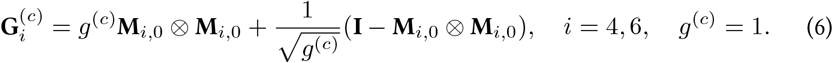

Assuming isochoric deformation:

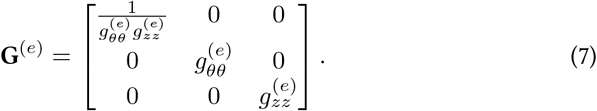

In the current study, the axial deposition stretch 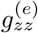 is derived from literature, as function of the patient’s age [45], while the circumferential deposition stretch 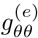 is calculated based on the condition of static equilibrium of the geometry experiencing the internal pressure at diastole [46]. For a given **s**, *H*_*dias*_, *p*_*dias*_ and **F** = **I**, and using the nonlinear equation solver ‘fsolve’ in Matlab R2019a, 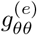 can be solved from:

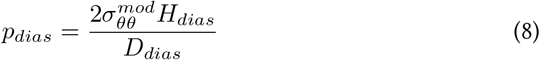

### Parameter fitting

The material parameters **s** and diastolic thickness *H*_*dias*_ are found by minimizing the objective *O* in equation 9.

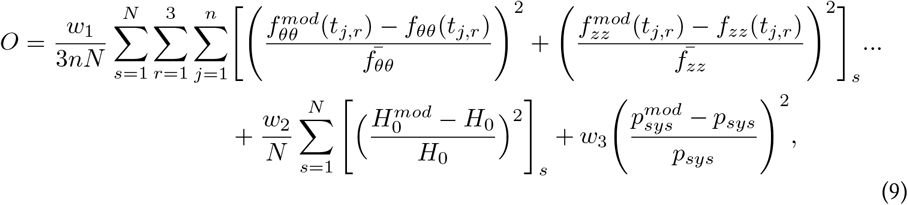

with 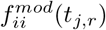 and *f*_*ii*_(*t*_*j,r*_) the model and experimental forces in the direction *i* = *θ, z* during the PB experiment at time point *t*_*j,r*_. *j* determines the index of the loading cycle at ratio *r, n* the total number of time points of a loading cycle, *s* the sample index, and *N* the total number of samples excised from the aneurysm. The weights *w*_1_, *w*_2_ and *w*_3_ were set to 0.3, 0.2 and 0.3, respectively. 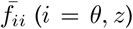 in equation 9 indicate the mean of all experimental forces along the *i*th direction.

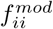 is calculated as:

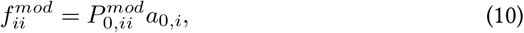

with *a*_0,*i*_ the cross-sectional area in the zero-stress state with its normal along *i*th direction. *P*^*mod*^ is the first Piola-Kirchhoff stress along *i*th direction pulled back to the zero-stress state and is calculated according to:

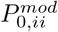 is calculated as:

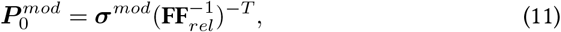

with **F** = **F**_*biax*_**F**_*rel*_, where **F**_*biax*_ is the deformation between the zero-stress state and the biaxially loaded state, and **F**_*rel*_ the deformation between the reference and the zero-stress state, see Figure 6.

To calculate 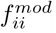, one should identify **F**_*rel*_ and **F**_*biax*_. Once the diastolic reference configuration is prestressed using **G**^(*c*)^ and **G**^(*e*)^, **F**_*rel*_, defined as

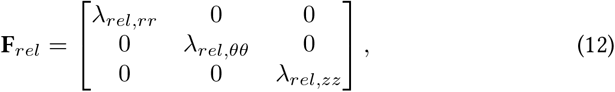

can be derived from the condition that 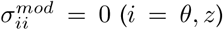. Assuming isochoric deformation, *i.e*. 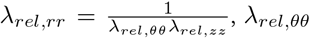 and *λ*_*rel,zz*_ are calculated using the nonlinear equation solver ‘fsolve’ in Matlab R2019a. Knowing **F**_*rel*_, the model thickness in the zero-stress state is then calculated as 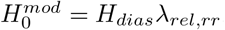.

**F**_*biax*_ = **F**_*LC*_**F**_*p*_, where **F**_*LC*_ is the deformation during the loading cycle and **F**_*p*_ the deformation caused by the preloading of the sample prior to the loading cycle, see [24]. No shear deformation during biaxial testing is assumed:

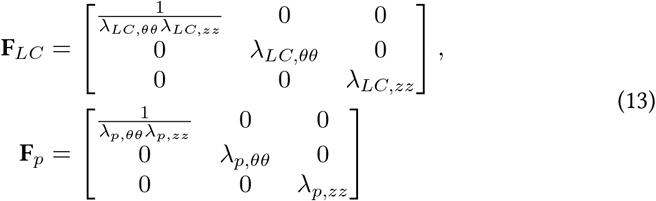

*λ*_*LC,θθ*_ and *λ*_*LC,zz*_ are experimentally measured by performing DIC between a biaxially loaded state and the preloaded state. *λ*_*p,θθ*_ and *λ*_*p,zz*_ are derived from the condition that 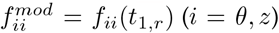 at each ratio *r*, using the nonlinear equation solver ‘fsolve’ in Matlab R2019a [24].

*a*_0,*j*_, used in equation 10, is determined as [24]:

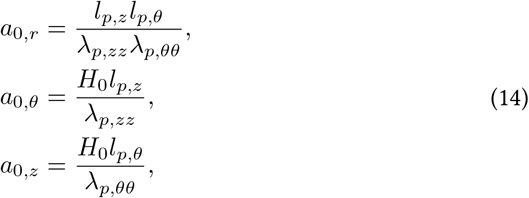

with *l*_*p,z*_ and *l*_*p,θ*_ the rake-to-rake distance in the preloaded state in the axial and circumferential direction, respectively.

The Cauchy stress at systole 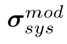 is calculated using equations 4 and 5, for a given **s, G**^(*c*)^, **G**^(*e*)^ and **F** = **F**_*sys*_, with

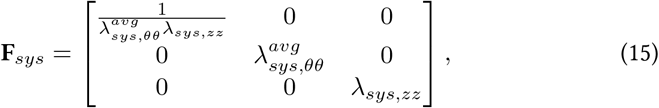

where 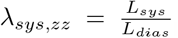 Assuming an incompressible extension-inflation between diastole and systole[4.6]

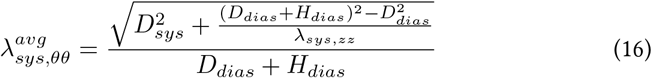

The model intraluminal systolic pressure is then: 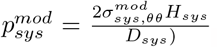, with 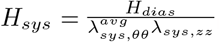

Equation 9 was minimized using the nonlinear least square solver in Matlab R2019a. 20 different start points are used to find a global minimum. The imposed boundaries on the parameters of **s** and *H*_*dias*_ are shown in Table 3. Because of the different ranges of the boundaries, the parameters were first scaled when performing the fitting. The fitting quality measures are defined through the use of the root mean squared error (RMSE):

**Table 3:**
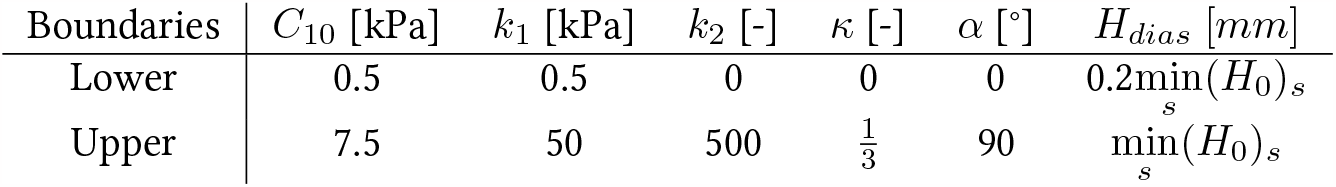
Boundaries of the material properties and *in vivo* thickness. *s* indicates the sample index.

**Table 4:** Overview of the fitted parameters, fitted *in vivo* diastolic thickness and fitting quality measures 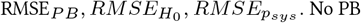. No PB test was performed for ATAA 6, 13 and 14.

| ATAA | $H_{dias}$<br>[mm] | $C_{10}$<br>[kPa] | $k_1$<br>[kPa] | $k_2$<br>[-] | $\kappa$<br>[-] | $\alpha$<br>[°] | $RMSE_{PB}$<br>[N] | $RMSE_{H_0}$<br>[mm] | $RMSE_{p_{sys}}$<br>[mmHg] |
| --- | --- | --- | --- | --- | --- | --- | --- | --- | --- |
| 1 | 2.66 | 2.90 | 820.50 | 165.58 | 0.22 | 57.83 | 0.27 | 0.83 | 0.16 |
| 2 | 2.07 | 19.80 | 74.70 | 124.67 | 0.23 | 33.69 | 0.22 | 0.19 | 0.28 |
| 3 | 1.73 | 21.40 | 203.40 | 231.31 | 0.22 | 22.37 | 0.10 | 0.20 | 0.14 |
| 4 | 1.90 | 27.30 | 2.00 | 232.18 | 0.13 | 12.19 | 0.25 | 0.39 | 0.12 |
| 5 | 1.64 | 19.20 | 91.00 | 71.06 | 0.17 | 33.91 | 0.11 | 0.29 | 0.11 |
| 7 | 2.09 | 31.50 | 17.50 | 157.93 | 0.15 | 1.90 | 0.28 | 0.52 | 0.15 |
| 8 | 1.70 | 19.90 | 334.50 | 33.92 | 0.17 | 32.94 | 0.11 | 0.54 | 0.03 |
| 9 | 3.31 | 13.60 | 54.90 | 352.63 | 0.21 | 38.92 | 0.18 | 0.43 | 0.03 |
| 10 | 2.38 | 14.00 | 128.00 | 68.42 | 0.17 | 30.55 | 0.08 | 0.54 | 0.10 |
| 11 | 1.51 | 17.50 | 125.60 | 52.52 | 0.16 | 7.10 | 0.06 | 0.17 | 0.02 |
| 12 | 3.03 | 16.00 | 171.10 | 53.85 | 0.20 | 15.78 | 0.12 | 0.31 | 0.01 |
| 15 | 2.13 | 0.80 | 72.00 | 13.85 | 0.22 | 32.88 | 0.05 | 0.10 | 0.04 |
| 16 | 2.11 | 14.10 | 296.00 | 77.69 | 0.20 | 33.36 | 0.05 | 0.52 | 0.08 |
| 17 | 2.08 | 14.80 | 73.40 | 35.65 | 0.18 | 3.66 | 0.10 | 0.56 | 0.01 |
| 18 | 2.48 | 9.10 | 170.10 | 160.73 | 0.21 | 37.87 | 0.07 | 0.49 | 0.05 |
| 19 | 1.47 | 14.90 | 272.50 | 19.71 | 0.15 | 40.33 | 0.04 | 0.16 | 0.08 |
| 20 | 1.64 | 12.80 | 81.20 | 33.66 | 0.13 | 3.72 | 0.14 | 1.57 | 0.01 |
| 21 | 1.92 | 10.00 | 178.70 | 103.40 | 0.18 | 37.13 | 0.15 | 0.84 | 0.08 |
| 22 | 2.08 | 12.00 | 277.40 | 201.60 | 0.23 | 27.00 | 0.06 | 0.41 | 0.05 |
| 23 | 2.79 | 12.40 | 5.70 | 256.38 | 0.16 | 1.44 | 0.08 | 0.42 | 0.04 |
| 24 | 1.14 | 8.40 | 456.90 | 4.05 | 0.14 | 43.16 | 0.10 | 0.26 | 0.02 |
| 25 | 1.63 | 13.70 | 51.10 | 24.44 | 0.17 | 5.77 | 0.07 | 0.41 | 0.03 |
| 26 | 1.26 | 18.20 | 259.50 | 0.72 | 0.16 | 0.29 | 0.06 | 0.44 | 0.00 |
| 27 | 1.24 | 11.10 | 69.20 | 22.99 | 0.10 | 0.24 | 0.05 | 0.34 | 0.04 |
| 28 | 1.59 | 16.20 | 434.90 | 25.06 | 0.19 | 35.37 | 0.03 | 0.37 | 0.03 |
| 29 | 1.03 | 13.80 | 22.10 | 12.88 | 0.12 | 2.60 | 0.04 | 0.25 | 0.02 |
| 30 | 1.73 | 9.30 | 193.20 | 33.38 | 0.17 | 7.56 | 0.05 | 0.58 | 0.07 |
| 31 | 1.73 | 12.60 | 122.50 | 17.18 | 0.20 | 0.81 | 0.03 | 0.35 | 0.00 |
| 32 | 1.66 | 15.00 | 372.60 | 34.23 | 0.21 | 0.21 | 0.04 | 0.59 | 0.07 |
| 33 | 1.71 | 15.40 | 255.50 | 13.47 | 0.18 | 19.25 | 0.09 | 0.49 | 0.03 |

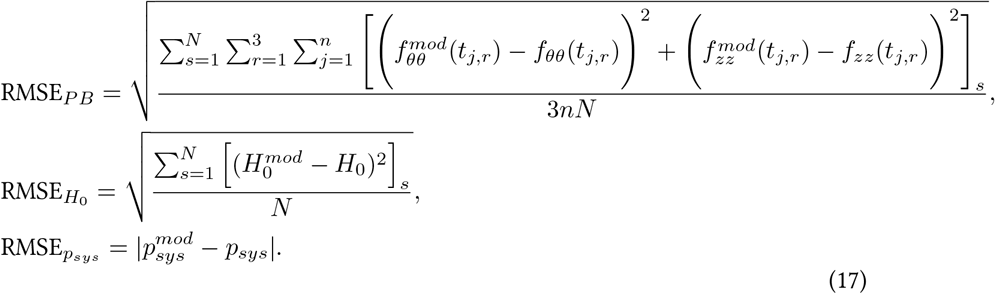

Once the parameters are fitted, the maximal first Piola-Kirchhoff stress at systole referring to the zero-stress state 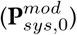 is calculated according to equation 11, with 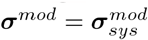 and **F** = **F**_*sys*_. 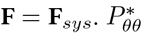 and 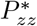 are defined as the circumferential and axial component of 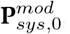, respectively.

Appendix

C: Fitted material parameters

## Notes

### Competing Interest Statement

The authors have declared no competing interest.

### Clinical Trial

NCT03142074

### Funding Statement

Research Foundation - Flanders (FWO) (Grants no: SB1SA9119N, SB1S56317) KU Leuven (C2-ADAPT)

### Author Declarations

The study was approved by the Ethics Committee Research UZ/KU Leuven (NCT03142074) and patient informed consent was obtained.

## References

[1] E. M. Isselbacher, Thoracic and abdominal aortic aneurysms, Circulation 111 (6) (2005) 816–828. doi:10.1161/01.CIR.0000154569.08857.7A.

[2] J. D. Humphrey, M. A. Schwartz, G. Tellides, D. M. Milewicz, Role of mechan-otransduction in vascular biology: Focus on thoracic aortic aneurysms and dis-sections, Circulation Research 116 (8) (2015) 1448–1461. doi:10.1161/CIRCRESAHA.114.304936.

[3] N. D. Anfinogenova, V. E. Sinitsyn, B. N. Kozlov, D. S. Panfilov, S. V. Popov, A. V. Vrublevsky, A. Chernyavsky, T. Bergen, V. V. Khovrin, W. Y. Ussov, Ex-isting and Emerging Approaches to Risk Assessment in Patients with Ascending Thoracic Aortic Dilatation, Journal of Imaging 8 (10) (2022). doi:10.3390/jimaging8100280.

[4] E. M. Isselbacher, O. Preventza, J. H. Black, J. G. Augoustides, A. W. Beck, M. A. Bolen, A. C. Braverman, B. E. Bray, M. M. Brown-Zimmerman, E. P. Chen, T. J. Collins, A. DeAnda, C. L. Fanola, L. N. Girardi, C. W. Hicks, D. S. Hui, W. S. Jones,V. Kalahasti, K. M. Kim, D. M. Milewicz, G. S. Oderich, L. Ogbechie, S. B. Promes,E. G. Ross, M. L. Schermerhorn, S. S. Times, E. E. Tseng, G. J. Wang, Y. J. Woo, 2022 ACC/AHA Guideline for the Diagnosis and Management of Aortic Disease: A Report of the American Heart Association/American College of Cardiology Joint Committee on Clinical Practice Guidelines, Circulation (dec 2022). doi: 10.1161/cir.0000000000001106.

[5] L. F. Hiratzka, G. L. Bakris, J. A. Beckman, R. M. Bersin, V. F. Carr, D. E. Casey, K. A. Eagle, L. K. Hermann, E. M. Isselbacher, E. A. Kazerooni, N. T. Kouchoukos,B. W. Lytle, D. M. Milewicz, D. L. Reich, S. Sen, J. A. Shinn, L. G. Svensson, D. M. Williams, A. K. Jacobs, S. C. Smith, J. L. Anderson, C. D. Adams, C. E. Buller, M. A. Creager, S. M. Ettinger, R. A. Guyton, J. L. Halperin, S. A. Hunt, H. M. Krumholz, F. G. Kushner, R. Nishimura, R. L. Page, B. Riegel, W. G. Stevenson, L. G. Tarkington, C. W. Yancy, 2010 ACCF/AHA/AATS/ACR/ASA/SCA/SCAI/SIR/STS/SVM guidelines for the diagnosis and management of patients with thoracic aortic disease: Executive summary: A report of the american college of cardiology foundation/american heart association task force on pra, Circulation 121 (13) (2010) 1544–1579. doi:10.1161/CIR.0b013e3181d47d48.

[6] S. Sherifova, G. A. Holzapfel, Biomechanics of aortic wall failure with a focus on dissection and aneurysm: A review, Acta Biomaterialia 99 (2019) 1–17. doi: 10.1016/j.actbio.2019.08.017. URL https://doi.org/10.1016/j.actbio.2019.08.017

[7] L. A. Pape, T. T. Tsai, E. M. Isselbacher, J. K. Oh, P. T. O’Gara, A. Evangelista,≤R. Fattori, G. Meinhardt, S. Trimarchi, E. Bossone, T. Suzuki, J. V. Cooper, J. B. Froehlich, C. A. Nienaber, K. A. Eagle, Aortic diameter ≤5.5 cm is not a good predictor of type A aortic dissection: Observations from the International Registry of Acute Aortic Dissection (IRAD), Circulation 116 (10) (2007) 1120–1127. doi:10.1161/CIRCULATIONAHA.107.702720.

[8] N. Kontopodis, D. Pantidis, A. Dedes, N. Daskalakis, C. V. Ioannou, The – Not So – Solid 5.5 cm Threshold for Abdominal Aortic Aneurysm Repair: Facts, Misinterpretations, and Future Directions, Frontiers in Surgery 3 (2016) 1. doi:10.3389/fsurg.2016.00001. URL http://www.ncbi.nlm.nih.gov/pmc/articles/PMC4725249/

[9] R. R. Davies, A. Gallo, M. A. Coady, G. Tellides, D. M. Botta, B. Burke, M. P. Coe, G. S. Kopf, J. A. Elefteriades, Novel measurement of relative aortic size predicts rupture of thoracic aortic aneurysms., The Annals of thoracic surgery 81 (1) (2006) 169–177. doi:10.1016/j.athoracsur.2005.06.026.

[10] M. A. Zafar, Y. Li, J. A. Rizzo, P. Charilaou, A. Saeyeldin, C. A. Velasquez, A. M. Mansour, S. U. Bin Mahmood, W. G. Ma, A. J. Brownstein, M. Tranquilli, J. Dum-farth, P. Theodoropoulos, K. Thombre, M. Tanweer, Y. Erben, S. Peterss, B. A. Ziganshin, J. A. Elefteriades, Height alone, rather than body surface area, suffices for risk estimation in ascending aortic aneurysm, Journal of Thoracic and Cardiovascular Surgery 155 (5) (2018) 1938–1950. doi:10.1016/j.jtcvs.2017.10.140. URL https://doi.org/10.1016/j.jtcvs.2017.10.140

[11] J. Wu, M. A. Zafar, Y. Li, A. Saeyeldin, Y. Huang, R. Zhao, J. Qiu, M. Tanweer,M. Abdelbaky, A. Gryaznov, J. Buntin, B. A. Ziganshin, S. K. Mukherjee, J. A. Rizzo, C. Yu, J. A. Elefteriades, Ascending Aortic Length and Risk of Aortic Adverse Events: The Neglected Dimension, Journal of the American College of Cardiology 74 (15) (2019) 1883–1894. doi:10.1016/j.jacc.2019.07.078.

[12] A. Duprey, O. Trabelsi, M. Vola, J. P. Favre, S. Avril, Biaxial rupture properties of ascending thoracic aortic aneurysms, Acta Biomaterialia 42 (2016) 273–285. doi:10.1016/j.actbio.2016.06.028. URL http://dx.doi.org/10.1016/j.actbio.2016.06.028

[13] C. Martin, W. Sun, T. Pham, J. Elefteriades, Predictive biomechanical analysis of ascending aortic aneurysm rupture potential, Acta biomaterialia 9 (12) (2013) 10.1016/j.actbio.2013.07.044. doi:10.1016/j.actbio.2013.07.044. URL http://www.ncbi.nlm.nih.gov/pmc/articles/PMC3872822/

[14] B. A. Ziganshin, M. A. Zafar, J. A. Elefteriades, Descending threshold for ascending aortic aneurysmectomy : Is it time for a “left-shift” in guidelines?, The Journal of Thoracic and Cardiovascular Surgery 157 (1) (2018) 37–42.doi:10.1016/j.jtcvs.2018.07.114. URL https://doi.org/10.1016/j.jtcvs.2018.07.114

[15] Y. J. Li, W. G. Ma, Y. Qi, J. M. Zhu, Y. Yang, L. Z. Sun, Does the 45 mm Size Cutoff for Ascending Aortic Replacement Predict Better Early Outcomes in Bicuspid Aortic Valve?, The Thoracic and Cardiovascular Surgeon 70 (4) (2022) 289–296. doi:10.1055/s-0040-1722197.

[16] J. A. Elefteriades, J. A. Rizzo, M. A. Zafar, B. A. Ziganshin, Jo ur na of, The Journal of Thoracic and Cardiovascular Surgery (2022). doi:10.1016/j.jtcvs.2022.07.033. URL https://doi.org/10.1016/j.jtcvs.2022.07.033

[17] M. K. Halushka, A. Angelini, G. Bartoloni, C. Basso, L. Batoroeva, P. Bruneval,L. M. Buja, J. Butany, G. D’Amati, J. T. Fallon, P. J. Gallagher, A. C. Gittenberger-De Groot, R. H. Gouveia, I. Kholova, K. L. Kelly, O. Leone, S. H. Litovsky, J. J. Maleszewski, D. V. Miller, R. N. Mitchell, S. D. Preston, A. Pucci, S. J. Radio,E. R. Rodriguez, M. N. Sheppard, J. R. Stone, S. K. Suvarna, C. D. Tan, G. Thiene,J. P. Veinot, A. C. Van Der Wal, Consensus statement on surgical pathology of the aorta from the Society for Cardiovascular Pathology and the Association forEuropean Cardiovascular Pathology: II. Noninflammatory degenerative diseases-Nomenclature and diagnostic criteria, Cardiovascular Pathology 25 (3) (2016) 247–257. doi:10.1016/j.carpath.2016.03.002.

[18] L. R. Bons, A. L. Duijnhouwer, S. Boccalini, A. T. van den Hoven, M. J. van der Vlugt, R. G. Chelu, J. S. McGhie, I. Kardys, A. E. van den Bosch, H. M. J. Siebelink, K. Nieman, A. Hirsch, C. S. Broberg, R. P. Budde, J. W. Roos-Hesselink, Intermodality variation of aortic dimensions: How, where and when to measure the ascending aorta, International Journal of Cardiology 276 (2019) 230–235. doi:10.1016/j.ijcard.2018.08.067. URL https://doi.org/10.1016/j.ijcard.2018.08.067

[19] R. Mosteller, Simplified Calculation of Body-Surface Area, New England Journal of Medicine 317 (17) (1987) 1098–1098. doi:10.1056/NEJM198710223171717. URL https://doi.org/10.1056/NEJM198710223171717 http://www.nejm.org/doi/abs/10.1056/NEJM198710223171717

[20] G. Koullias, R. Modak, M. Tranquilli, D. P. Korkolis, P. Barash, J. A. Elefteriades, Mechanical deterioration underlies malignant behavior of aneurysmal human ascending aorta, The Journal of thoracic and cardiovascular surgery 130 (3) (2005) 677.e1–677.e9. doi:10.1016/J.JTCVS.2005.02.052. URL https://pubmed.ncbi.nlm.nih.gov/16153912/

[21] M. Smoljkić, H. Fehervary, P. Van den Bergh, A. Jorge-Pen∼as, L. Kluyskens,S. Dymarkowski, P. Verbrugghe, B. Meuris, J. Vander Sloten, N. Famaey, Biomechanical Characterization of Ascending Aortic Aneurysms, Biomechanics and Modeling in Mechanobiology 16 (2) (2017) 705–720. doi:10.1007/s10237-016-0848-4.

[22] C. Martin, W. Sun, C. Primiano, R. McKay, J. Elefteriades, Age-dependent ascending aorta mechanics assessed through multiphase CT, Annals of Biomedical Engineering 41 (12) (2013) 2565–2574. doi:10.1007/s10439-013-0856-9.

[23] L. Maes, H. Fehervary, J. Vastmans, S. J. Mousavi, S. Avril, N. Famaey, Constrained mixture modeling affects material parameter identification from planar biaxial tests, Journal of the Mechanical Behavior of Biomedical Materials 95 (March) (2019) 124–135. doi:10.1016/j.jmbbm.2019.03.029.

[24] K. Vander Linden, H. Fehervary, L. Maes, N. Famaey, An improved parameter fitting approach of a planar biaxial test including the experimental prestretch, Journal of the Mechanical Behavior of Biomedical Materials 34 (2022) 105389. doi:10.1016/J.JMBBM.2022.105389.

[25] A. Masri, V. Kalahasti, L. G. Svensson, E. E. Roselli, D. Johnston, D. Hammer, P. Schoenhagen, B. P. Griffin, M. Y. Desai, Aortic Cross-Sectional Area/Height Ratio and Outcomes in Patients with a Trileaflet Aortic Valve and a Dilated Aorta, Circulation 134 (22) (2016) 1724–1737.doi:10.1161/CIRCULATIONAHA.116.022995. URL https://www.ahajournals.org/doi/abs/10.1161/CIRCULATIONAHA.116.022995

[26] J. Z. Goldfinger, J. L. Halperin, M. L. Marin, A. S. Stewart, K. A. Eagle, V. Fuster, Thoracic aortic aneurysm and dissection., Journal of the American College of Cardiology 64 (16) (2014) 1725–1739. doi:10.1016/j.jacc.2014.08.025.

[27] S. Polzer, J. KracÍk, T. Novotný L. Kubýček, R. Staffa, M. L. Raghavan, Methodol-ogy for Estimation of Annual Risk of Rupture for Abdominal Aortic Aneurysm, Computer Methods and Programs in Biomedicine 200 (2021). doi:10.1016/j.cmpb.2020.105916.

[28] D. P. Howard, A. Banerjee, J. F. Fairhead, A. Handa, L. E. Silver, P. M. Rothwell, Age-specific incidence, risk factors and outcome of acute abdominal aortic aneurysms in a defined population, British Journal of Surgery 102 (8) (2015) 907–915. doi:10.1002/bjs.9838.

[29] J. A. Elefteriades, S. K. Mukherjee, H. Mojibian, Discrepancies in Measurement of the Thoracic Aorta: JACC Review Topic of the Week, Journal of the American College of Cardiology 76 (2) (2020) 201–217. doi:10.1016/j.jacc.2020.03.084.

[30] A. Zierer, S. J. Melby, J. G. Lubahn, G. A. Sicard, R. J. Damiano, M. R. Moon, Elective Surgery for Thoracic Aortic Aneurysms: Late Functional Status and Quality of Life, The Annals of Thoracic Surgery 82 (2) (2006) 573–578. doi: 10.1016/J.ATHORACSUR.2006.03.042.

[31] M. H. Heusinkveld, S. Quicken, R. J. Holtackers, W. Huberts, K. D. Reesink,T. Delhaas, B. Spronck, Uncertainty quantification and sensitivity analysis of an arterial wall mechanics model for evaluation of vascular drug therapies, Biomechanics and Modeling in Mechanobiology 17 (1) (2018) 55–69. doi: 10.1007/s10237-017-0944-0.

[32] L. Bruder, J. Pelisek, H. H. Eckstein, M. W. Gee, Biomechanical rupture risk assessment of abdominal aortic aneurysms using clinical data: A patient-specific, probabilistic framework and comparative case-control study, PLoS ONE 15 (11 November) (2020). doi:10.1371/journal.pone.0242097. URL http://dx.doi.org/10.1371/journal.pone.0242097

[33] P. Hauseux, J. S. Hale, S. Cotin, S. P. Bordas, Quantifying the uncertainty in a hyperelastic soft tissue model with stochastic parameters, Applied Mathematical Modelling 62 (2018) 86–102. doi:10.1016/j.apm.2018.04.021. URL https://doi.org/10.1016/j.apm.2018.04.021

[34] S. Polzer, T. C. Gasser, Biomechanical rupture risk assessment of abdominal aortic aneurysms based on a novel probabilistic rupture risk index, Journal of the Royal Society Interface 12 (113) (2015). doi:10.1098/rsif.2015.0852.

[35] M. Liu, L. Liang, Q. Zou, Y. Ismail, X. Lou, G. Iannucci, E. P. Chen, B. G. Leshnower, J. A. Elefteriades, W. Sun, A probabilistic and anisotropic failure metric for ascending thoracic aortic aneurysm risk assessment, Journal of the Mechanics and Physics of Solids 155 (January) (2021). doi:10.1016/j.jmps.2021.104539.

[36] R. A. Macrae, K. Miller, B. J. Doyle, Methods in Mechanical Testing of Arterial Tissue: A Review, Strain 52 (5) (2016) 380–399. doi:10.1111/str.12183.

[37] A. Romo, P. Badel, A. Duprey, J. P. Favre, S. Avril, In vitro analysis of localized aneurysm rupture, Journal of Biomechanics 47 (3) (2014) 607–616. doi:10.1016/j.jbiomech.2013.12.012.

[38] S. Lin, M. C. Morgant, D. M. Mar ćn-Castrillón, P. M. Walker, L. S. Aho Gl élé,A. Boucher, B. Presles, O. Bouchot, A. Lalande, Aortic local biomechanical properties in ascending aortic aneurysms, Acta Biomaterialia 149 (2022) 40–50. doi:10.1016/j.actbio.2022.06.019.

[39] B. A. Alhafez, V. T. T. Truong, D. Ocazionez, S. Sohrabi, H. Sandhu, A. Estrera,H. J. Safi, A. Evangelista, L. D. S. Hurtado, A. Guala, S. K. Prakash, Aortic arch tortuosity, a novel biomarker for thoracic aortic disease, is increased in adults with bicuspid aortic valve, International Journal of Cardiology 284 (2019) 84–89. doi:10.1016/j.ijcard.2018.10.052. URL /pmc/articles/PMC6436988//pmc/articles/PMC6436988/?report=abstracthttps://www.ncbi.nlm.nih.gov/pmc/articles/PMC6436988/

[40] S. Farzaneh, O. Trabelsi, B. Chavent, S. Avril, Identifying Local Arterial Stiffness to Assess the Risk of Rupture of Ascending Thoracic Aortic Aneurysms, Annals of Biomedical Engineering 47 (4) (2019) 1038–1050. doi:10.1007/s10439-019-02204-5.

[41] W. A. Zoghbi, D. Adams, R. O. Bonow, M. Enriquez-Sarano, E. Foster, P. A. Grayburn, R. T. Hahn, Y. Han, J. Hung, R. M. Lang, S. H. Little, D. J. Shah, S. Shernan, P. Thavendiranathan, J. D. Thomas, N. J. Weissman, Recommendations for Noninvasive Evaluation of Native Valvular Regurgitation: A Report from the American Society of Echocardiography Developed in Collaboration with the Society for Cardiovascular Magnetic Resonance, Journal of the American Society of Echocardiography 30 (4) (2017) 303–371. doi:10.1016/j.echo.2017.01.007. URL http://dx.doi.org/10.1016/j.echo.2017.01.007

[42] M. S. Sacks, Biaxial mechanical evaluation of planar biological materials, Journal of Elasticity 61 (1-3) (2000) 199–246. doi:10.1023/A:1010917028671.

[43] T. C. Gasser, R. W. Ogden, G. Holzapfel, Hyperelastic modelling of arterial layers with distributed collagen fibre orientations, Journal Of The Royal Society Interface 3 (6) (2006) 15–35. doi:10.1098/rsif.2005.0073.

[44] C. J. Cyron, R. C. Aydin, J. D. Humphrey, A homogenized constrained mix-ture (and mechanical analog) model for growth and remodeling of soft tis-sue, Biomechanics and Modeling in Mechanobiology 15 (6) (2016) 1389–1403. doi:10.1007/s10237-016-0770-9.

[45] L. Horn Í, M. Netušil, T. Voňavková, Axial prestretch and circumferen-tial distensibility in biomechanics of abdominal aorta., Biomechanics and modeling in mechanobiology 13 (4) (2014) 783–799. doi:10.1007/s10237-013-0534-8.

[46] K. Vander Linden, M. Ghasemi, L. Maes, J. Vastmans, N. Famaey, Layer-specific fiber distribution in arterial tissue modeled as a constrained mixture, International Journal for Numerical Methods in Biomedical Engineering (2022) e3608doi:10.1002/CNM.3608.

